## Supplementary Figure for "Correlations in sleeping patterns and circadian preference between spouses"

**Supplementary Figures**

Supplementary Figure 1 – Participant flow diagram

488,377 genotyped participants

463,827 European descent

47,549 spouse pairs

47,420 pairs with self-reported sleep*

3,454 pairs with valid accelerometer-derived sleep*

*At least one measure

Supplementary Figure 2 – Cross-trait spousal phenotypic correlations in UK Biobank

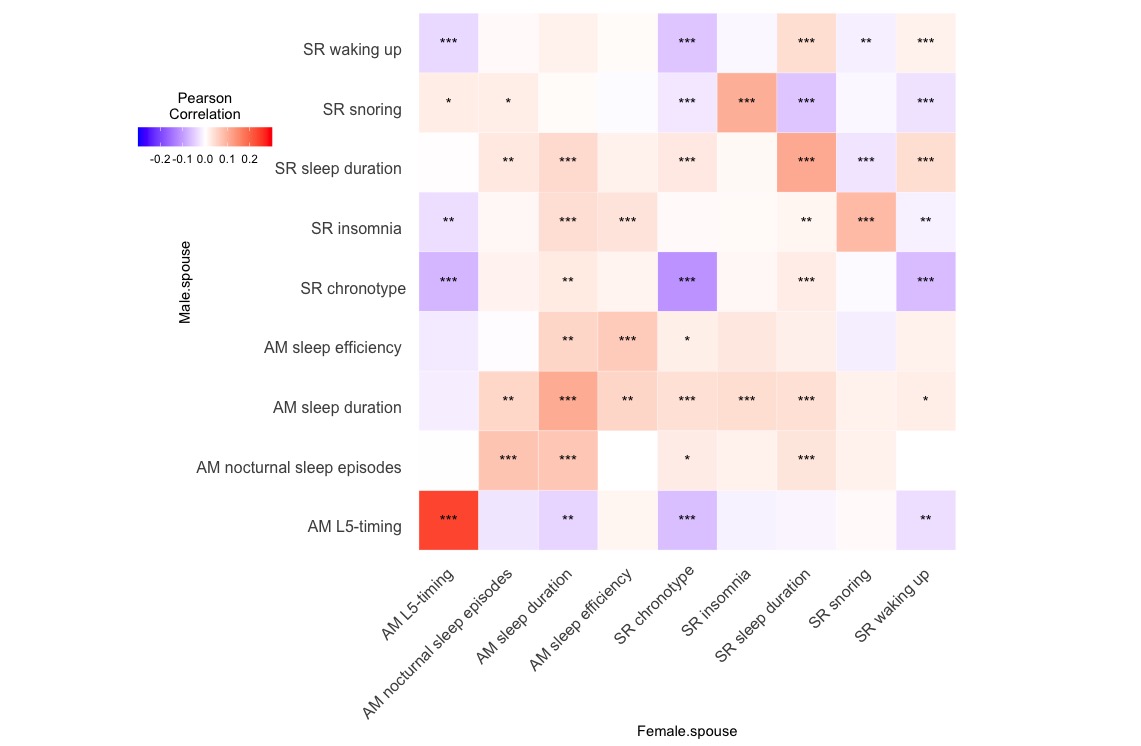

SR = self-reported; AM = accelerometer measured

Supplementary Figure 3 – Cross-trait spousal phenotypic correlations in 23andMe

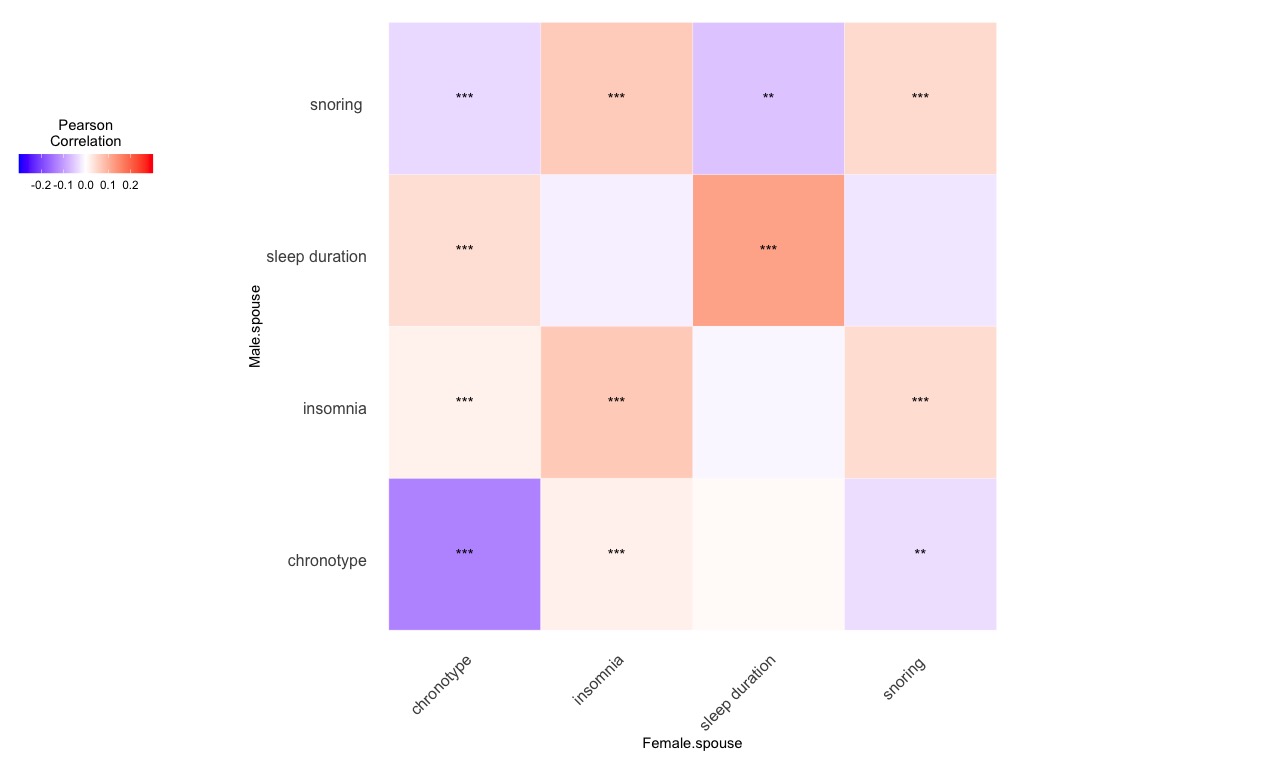

Supplementary Figure 4 - Comparison of causal estimates from Mendelian randomization between sexes in UK Biobank
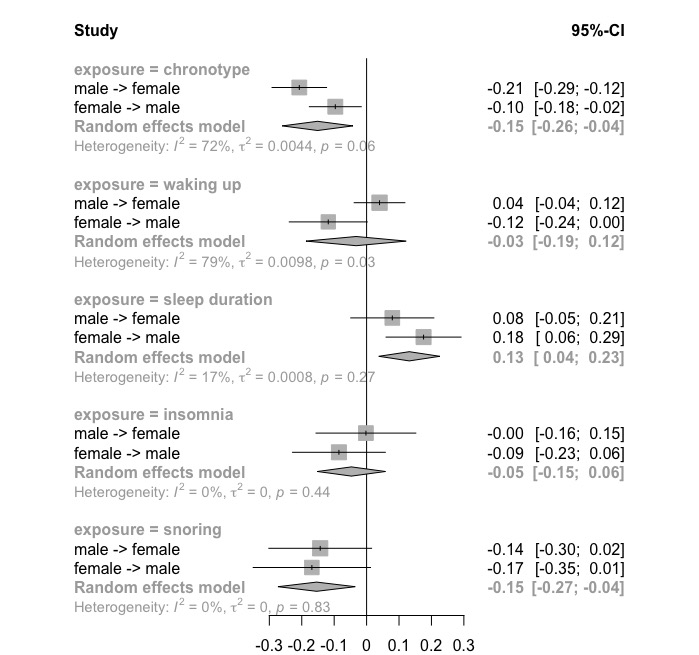

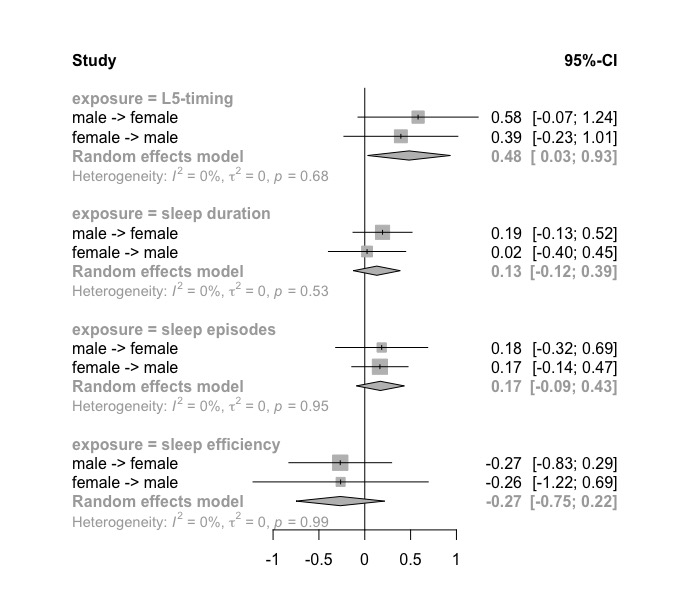

Supplementary Figure 5 - Cross-trait causal estimates from Mendelian randomization in UK Biobank

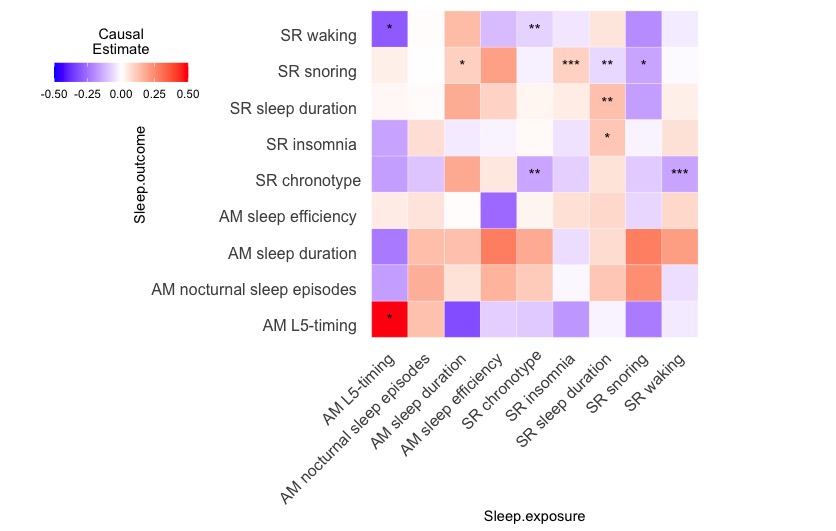

Supplementary Figure 6 - Spousal genotypic correlations between sleep traits in UK Biobank

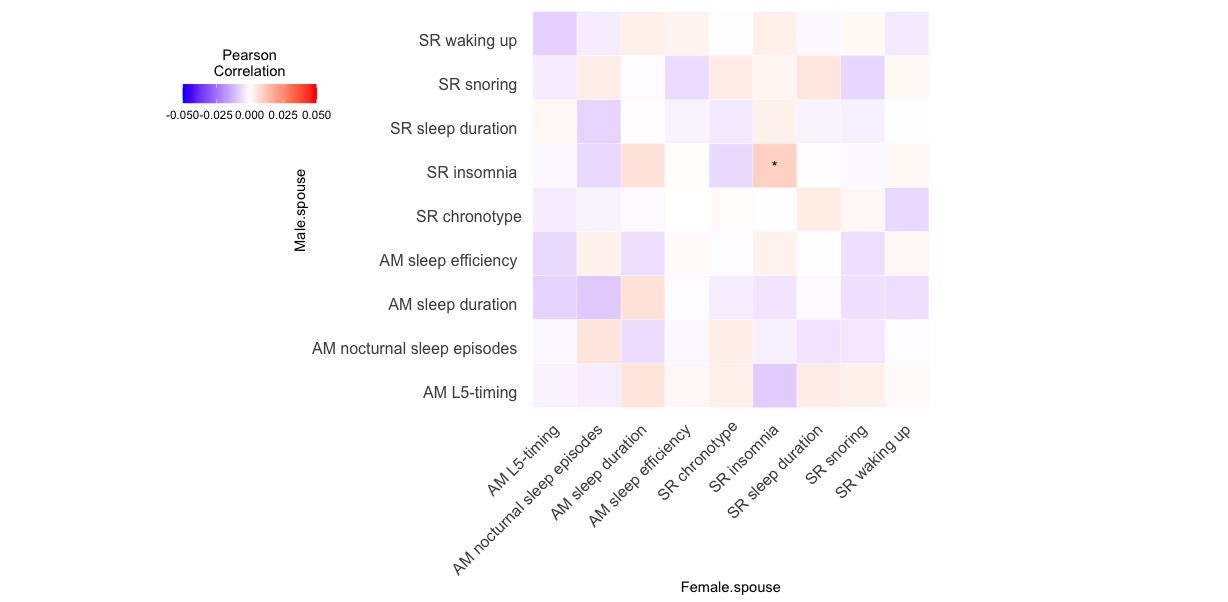

Supplementary Figure 7 – Genetic risk score correlations between spouses in UK Biobank

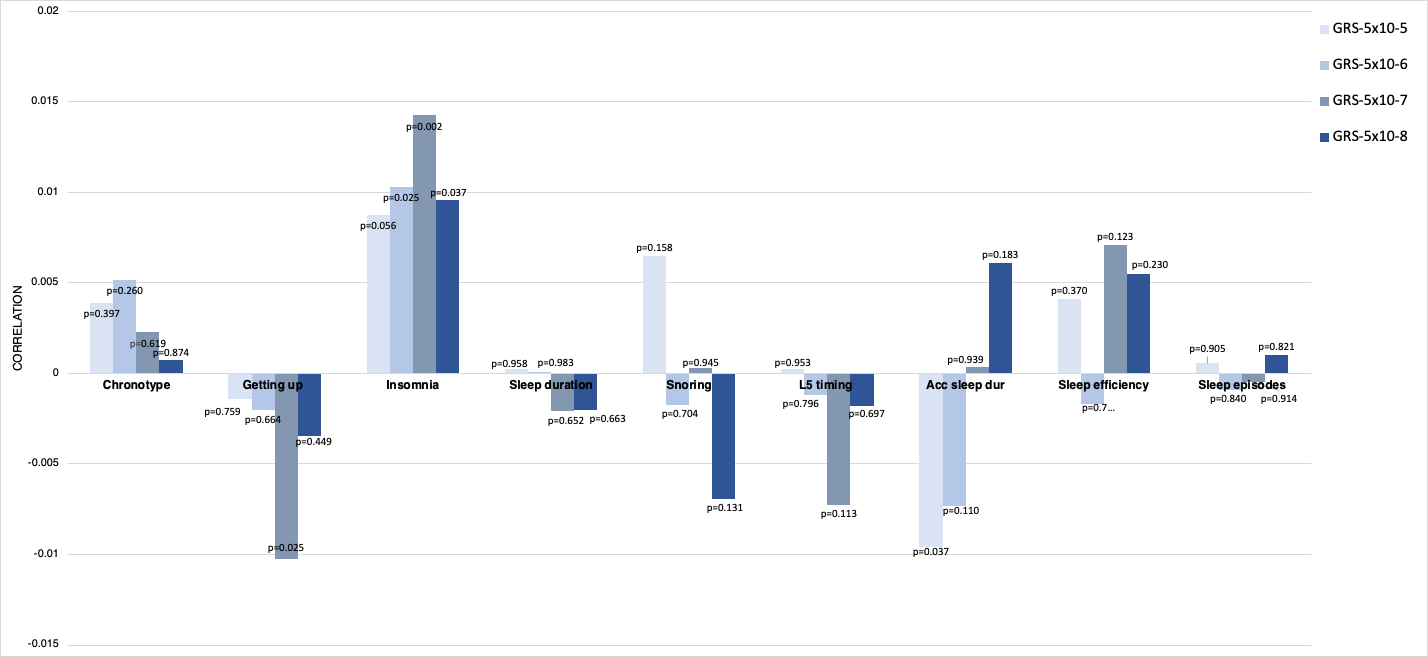

Supplementary Figure 8 - Difference in effects by age, time and location in UK Biobank

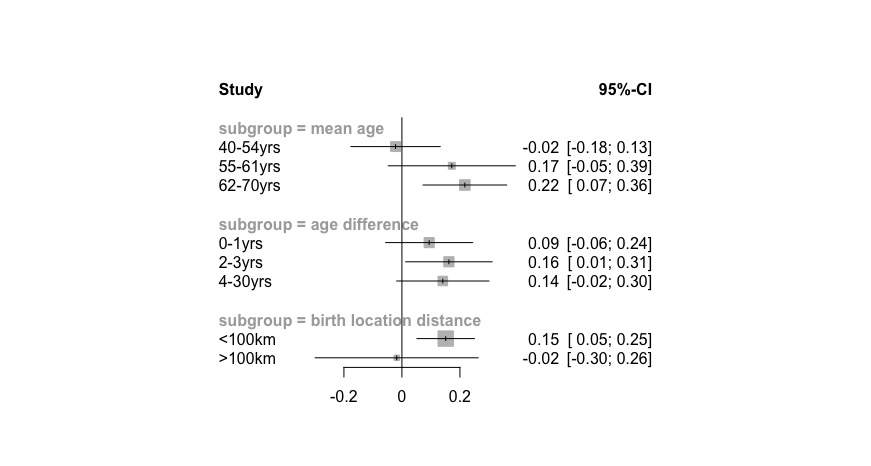

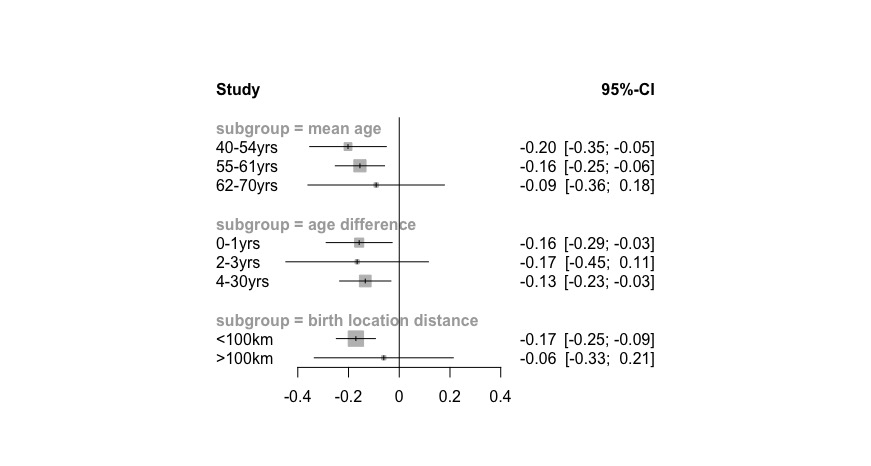

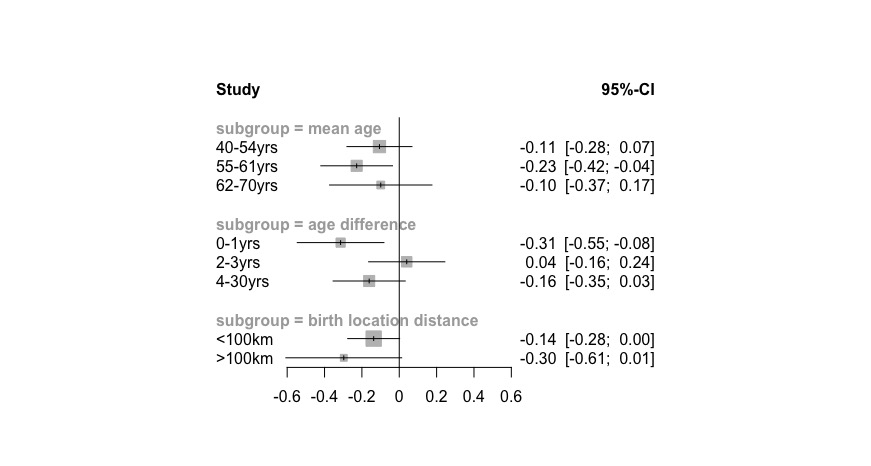

**Chronotype**

**Sleep duration**

**Snoring**

**L5-timing**

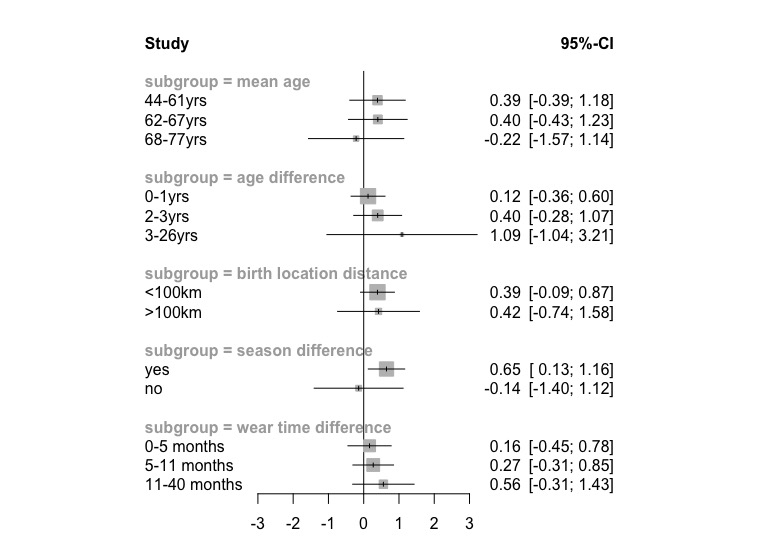
